# A study of metabolic syndrome in patients of HIV on ART in India

**DOI:** 10.1101/2020.12.28.20248924

**Authors:** V K Sashindran, Anchit Raj Singh

## Abstract

Metabolic syndrome is a risk factor for cardiovascular disease. With improvement in therapy for HIV, morbidity related to metabolic syndrome becomes a focus of interest. Change in nutritional status and introduction of newer regimens of ART are changing the impact of metabolic syndrome on patients of HIV. Few studies in recent times from developing countries have addressed this question. This is a cross sectional study to study the prevalence of Metabolic Syndrome in patients of HIV on ART from clinics in a tertiary care hospital in India. Data from 1208 patients of HIV on ART was analyzed and metabolic syndrome was seen in 257(21.3%) patients.

The high prevalence of metabolic syndrome in patients of HIV in India found in this study gives an insight on the morbidity of noncommunicable diseases in patients with HIV in India in recent times.

## Introduction

Metabolic Syndrome (MetS) is a group of factors that increase the risk of cardiovascular disease.^1^ MetS is defined as the presence of three or more of the following components: abdominal obesity, elevated triglycerides (Tg), reduced HDL cholesterol (HDL-c), elevated blood pressure (BP) and elevated fasting plasma glucose (FPG). The U.S. National Cholesterol Education Program Adult Treatment Panel III (NCEP ATP III) has the specified the values of these parameters as: abdominal obesity, waist circumference (WC) more than 40 inches in males and 35 inches in females, Tg >150mg/dL or drug treatment for elevated Tg, reduced HDL-c of < 40mg/dL in men and 50mg/dL in women, elevated BP >130/85mmHg or drug treatment for elevated BP, raised FPG > 100mg/dL or drug treatment for MetS.^2^

Several studies have not used the waist circumference and have adopted body mass index (BMI) cutoffs for MetS.^3^

Antiretroviral therapy (ART) regimes in the past had increased risk of lipodystrophy and the chronic inflammatory nature of the disease make patients of HIV more prone to MetS. Since ART regimens are changing, guidelines for starting ART have evolved and survival on therapy has increased, it has contributed to MetS being a major contributor to HIV related morbidity.^4^ The manifestation of MetS in a developing country like India needs to be studied in view of malnutrition in the background of high prevalence of opportunistic infections like tuberculosis.^4^

Estimates on prevalence of MetS in people living with HIV/AIDS (PLHA) have ranged from 7% to 30%. A recent meta-analysis of HIV and MetS calculated an overall prevalence of 16.7%.^5^

There have not been any adequately powered studies on metabolic syndrome in HIV in India in the recent past and with increasing life expectancy in patients with HIV, it is important to consider the morbidity of metabolic syndrome, the pathogenesis of which is closely associated with the chronic immune activation in HIV.^6^

## Materials and Methods

This is a cross sectional study carried out in a tertiary care hospital in India. Patients were recruited from two ART clinics in the hospital. Patients aged 18-60 years were included from Dec 2016 to Nov 2018. Pregnant women, patients with alcohol dependence syndrome and patients with suboptimal adherence to ART were excluded from the study. The NCEP ATP III definition of metabolic syndrome was used to identify patients with metabolic syndrome and BMI>25 was used in place of waist circumference on the lines of similar studies. Historical, clinical and laboratory parameters were recorded at the time of the visit. Institutional Ethics Committee Clearance was taken.

Data analysis was done using SPSS version 25. Prevalence was calculated in the population of interest. Chi square test was applied for rejection of the null hypothesis. Logistic regression was used for testing association. Patients with missing data related to parameters of metabolic syndrome were excluded.

## Resuts

In this study, 1208 patients were included in the final analysis after excluding patients according to the exclusion criteria. The baseline characteristics are detailed in Table 1.

**Table 1.** Baseline characteristics

| Characteristics | All patients (N=1208) |
| --- | --- |
| Age, Mean±SD -yr | 40.6 ± 9.3 |
| Sex, Male no (%) | 792 (65.6) |
| BMI, Mean±SD -kg/m<sup>2</sup> | 22.8 ± 3.9 |
| ART duration, Mean±SD -yr | 5.47 ± 3.86 |
| CD4 count, Mean±SD -cells/mm<sup>3</sup> | 437 ± 261 |
| Past Opportunistic Infections no(%) | 323(26.7) |

MetS was seen in 257 (21.3%) patients. The association between age and MetS revealed a significant (p<0.001) positive correlation (OR=1.0536; 95%CI=1.037-1.070).The association between MetS did not have any significant(p=0.505) relation with sex. ART duration and MetS was studied, which showed a statistically significant (p<0.001) positive correlation (OR=1.089; 95%CI=1.053-1.128). MetS had a significant association (p=0.03) with presence of past opportunistic infections

## Discussion

The prevalence of MetS in HIV patient on ART has been consistent with previous studies, although within range, on the higher side as compared to previous studies. Most studies use the ATP III definition, which might have a lower prevalence of MetS, therefore the prevalence with the IDF or WHO definition should be similar or higher. Also previous Indian studies have reported a prevalence of MetS in patients on ART of 20 and 25%.^7^ Although some Indian studies have found patients with HIV to be more malnourished compared to the West, the same was not seen in our study. There could be many explanations for this like: improvement in standards of living, economic development, early initiation of ART, reduction in stigma associated with HIV, inclusion of strata of society with better nutritional standards than the general population and expansion of state AIDS control societies and RNTCP acts. This study showed a statistically significant association between age and MetS. This finding is similar to prior studies.^8^ This study found no gender differences in MetS similar to previous studies.^9^ This study also found that the presence of OIs was associated with MetS, and might be significant even though previous studies have shown no such relation. It could be due to a state of chronic inflammation, exposure to greater duration and multiple lines of ART, however this needs to be studied further.

The large sample size, unlike other studies in Indian population is a strength of this study. This study also included patients from multiple geographical locations which are usually not seen in single center studies.

It was a cross sectional study, hence the inherent weakness of the study design, such as both exposures and outcomes were assessed at one point in time and it was difficult to establish a temporal relation between exposure and outcome were noted. There was no follow up of patients and this might have overestimated the prevalence of MetS compared to studies which used two observations separated in time to classify patients with MetS. This study was done at a point in time which was quite dynamic and in the preceding years had seen a constant change in treatment guidelines, which could have affected the data distribution.^10^

The findings of this study are limited to a single center and young and middle aged adults, with only cross sectional data being available. Therefore, before we generalize the results of this study, we will need to conduct it at multiple centers, with long term follow up to establish a temporal association. However, with the large sample size with population from all parts of India and multiple factors studied, we can be fairly confident that the findings of this study will be reproducible to a great extent.

## Data Availability

Requests to access the data should be addressed to

